# Recurrence patterns and adjuvant therapy in endometrioid early-stage endometrial cancer with a p53-abnormal pattern

**DOI:** 10.64898/2026.10.14.26365577

**Authors:** Sang hun Lee, Putri Deva Karimah, Kidong Kim, Se Ik Kim, Woo Yeon Hwang, Min Kyung Kim, Jae-Hoon Kim, Chel Hun Choi, Yoo-Kyung Lee

## Abstract

**Objective:** Current guidelines recommend different adjuvant treatment strategies for women with endometrioid early-stage endometrial cancer exhibiting a p53-abnormal pattern. We aimed to evaluate recurrence patterns and explore the clinical implications for adjuvant therapy in this population.

**Materials and methods:** This multicenter retrospective study included 86 patients with endometrioid, p53-abnormal, FIGO stage I–II endometrial cancer treated between 2013 and 2023 at six hospitals in the Republic of Korea. Clinicopathologic data were collected through medical record review. Progression-free survival (PFS) was estimated using the Kaplan– Meier method, and prognostic factors were analyzed using Cox proportional hazards regression models.

**Results:** The 12-, 24-, and 36-month PFS rates were 91%, 78%, and 76%, respectively. Among the 16 patients who experienced recurrence, 12 (75%) had distant metastases. Adjuvant chemotherapy showed a trend toward improved PFS compared with adjuvant radiation therapy with or without chemotherapy (p=0.10). In multivariable analysis, the p53-overexpression pattern was independently associated with worse PFS compared with the p53-null pattern (HR 8.59, 95% CI 1.61–45.79, p=0.01).

**Conclusion:** Given the poor PFS, the predominance of distant recurrence and the observed trend toward improved PFS with chemotherapy, adjuvant chemotherapy may be considered in women with endometrioid early-stage endometrial cancer exhibiting a p53-abnormal pattern. Further studies are warranted to clarify the prognostic impact of p53 abnormality subtypes.

## Introduction

Endometrial cancer represents a major global health concern, with a steadily increasing incidence in both high-income and low-to middle-income countries. In 2020, approximately 417,000 new cases were diagnosed worldwide, resulting in nearly 100,000 deaths [1–3]. Although early-stage disease is generally associated with favorable survival outcomes, recent nationwide analyses suggest that overall survival gains have been modest, particularly among patients with advanced-stage or high-risk subtypes [4–6].

Molecular classification has significantly refined prognostic stratification in endometrial cancer. The Cancer Genome Atlas (TCGA) classifies endometrial cancer into four molecular subtypes: POLE-ultramutated, mismatch repair–deficient (dMMR), TP53-abnormal (p53-abnormal), and no specific molecular profile (NSMP). These subtypes demonstrate distinct prognostic differences, with POLE-mutated tumors associated with excellent outcomes, p53-abnormal cancers showing the poorest survival, and dMMR and NSMP tumors exhibiting intermediate prognoses [5].

The incorporation of molecular classification into clinical practice varies across major guidelines. The European ESGO/ESTRO/ESP guidelines integrate molecular markers, including p53 status, into risk stratification and treatment recommendations [7]. In contrast, the National Comprehensive Cancer Network (NCCN) guidelines do not fully incorporate molecular classification into adjuvant treatment algorithms [8].

As a result, recommendations for adjuvant therapy differ between guidelines. For women with endometrioid early-stage endometrial cancer exhibiting a p53-abnormal pattern, the ESGO/ESTRO/ESP guidelines recommend adjuvant chemotherapy with or without radiation based on molecular risk stratification [7], whereas the NCCN guidelines generally recommend adjuvant radiation with or without chemotherapy [8].

Given these discrepancies, we aimed to evaluate recurrence patterns and explore the clinical implications for adjuvant therapy in women with endometrioid early-stage endometrial cancer with a p53-abnormal pattern.

## Materials and methods

### Study Design and Setting

This retrospective multicenter study was conducted between January 2013 and June 2023 at six tertiary hospitals in the Republic of Korea. The study protocol was approved by the Institutional Review Board of each participating institution (representative approval No. B-2310-856-101). The requirement for informed consent was waived owing to the retrospective study design.

The authors used ChatGPT (OpenAI, San Francisco, CA, USA) to assist with English language editing. The authors reviewed and revised the content to ensure its accuracy and take full responsibility for the final manuscript.

### Patient Selection

Eligible patients were women with newly diagnosed endometrioid-type endometrial cancer who underwent primary surgical treatment, including hysterectomy, between January 1, 2013, and June 30, 2023. Inclusion criteria were as follows: (1) histologically confirmed myometrial invasion; (2) International Federation of Gynecology and Obstetrics (FIGO) 2009 stage I or II disease; and (3) p53-abnormal status determined by immunohistochemistry (IHC). Patients were excluded if they had (1) received progestin therapy for endometrial cancer within 6 months prior to surgery or (2) undergone neoadjuvant chemotherapy or radiotherapy.

### Determination of p53 Status

The p53 status was retrieved from pathology reports in the medical records. IHC staining patterns were categorized as follows: (1) complete absence of nuclear staining with intact internal controls (null pattern); (2) diffuse, strong nuclear overexpression in >80% of tumor cells (overexpression pattern); (3) predominant cytoplasmic staining (cytoplasmic pattern); and (4) abnormal staining pattern not otherwise classifiable (NOS pattern). All of the above patterns were classified as p53-abnormal.

### Data Collection

Clinicopathologic data were extracted from medical records at each participating institution and compiled into a centralized, de-identified database. Collected variables included demographic and clinical characteristics (age, menopausal status, body mass index) and tumor-related factors (histologic grade, depth of myometrial invasion, lymphovascular space invasion [LVSI], lymph node metastasis, mismatch repair [MMR] status, estrogen receptor [ER] expression, and FIGO stage).

Treatment-related variables (adjuvant therapy modality, chemotherapy regimen, and number of cycles) and follow-up information (recurrence status, date and site of recurrence, survival status, and last follow-up date) were also recorded. However, complete chemotherapy data including the number of cycles and treatment intensity were not available for all patients. High-intermediate-risk criteria were defined according to the Gynecologic Oncology Group (GOG)-99 criteria [9].

The primary endpoint was progression-free survival (PFS), defined as the interval from the date of primary surgery to the date of first documented recurrence. Patients without recurrence were censored at the date of last follow-up. Recurrence sites were categorized as vaginal, regional, or distant based on radiologic and/or histologic confirmation.

### Statistical Analysis

PFS was estimated using the Kaplan–Meier method, and differences between groups were assessed using the log-rank test. Cox proportional hazards regression models were used to evaluate potential prognostic factors. Variables with p<0.05 in univariable analysis or those considered clinically relevant were included in the multivariable model.

Analyses were performed using Python (lifelines package, version 0.30.1). Missing data were handled using complete-case analysis. Patients with missing values for a given variable were excluded from analyses involving that variable. Multivariable analysis was conducted using cases with complete data for all included variables (n=67).

## Results

### Baseline Characteristics

The baseline characteristics of the 86 patients are summarized in Table 1. The median age was 58 years (range, 35–84), and 72% were postmenopausal. Most patients had FIGO stage IA disease (70%), histologic grade 2 tumors (37%), and <50% myometrial invasion (73%). MMR status was available for 38 patients, of whom 17 (45%) demonstrated MMR deficiency. The most common p53-abnormal pattern was the null type (69%), followed by overexpression (27%) and other abnormal patterns (5%). Thirty patients (35%) met the high-intermediate-risk criteria. Adjuvant therapy was administered to 40% of patients, most commonly chemotherapy alone (13%) or vaginal brachytherapy (12%).

**Table 1.** Baseline characteristics (n=86)

| Variable | Category | Value |
| --- | --- | --- |
| Age, year, mean ± SD | | 58 ± 10 |
| BMI, mean ± SD | | 26 ± 5 |
| Menopause, n (%) | Premenopause | 24 (28) |
|  | Postmenopause | 62 (72) |
| Stage, FIGO 2009, n (%) | IA | 60 (70) |
|  | IB | 19 (22) |
|  | II | 7 (8) |
| Histologic grade, n (%) | 1 | 30 (35) |
|  | 2 | 32 (37) |
|  | 3 | 23 (27) |
|  | Unknown | 1 (1) |
| Depth of myometrial invasion, n (%) | <1/2 | 63 (73) |
| | ≥1/2 | 20 (23) |
|  | Unknown | 3 (3) |
| Lymphovascular space invasion, n (%) | Absent | 72 (84) |
|  | Present | 14 (16) |
| MMR status, n (%) | Proficient | 21 (24) |
|  | Deficient | 17 (20) |
|  | Unknown | 48 (56) |
| p53 abnormality subtype, n (%) | Null | 59 (69) |
|  | Overexpression | 23 (27) |
|  | Others | 4 (5) |
| ER expression, n (%) | Positive | 62 (72) |
|  | Negative | 13 (15) |
|  | Unknown | 11 (13) |
| High-intermediate risk criteria, n (%) | Not satisfying | 54 (63) |
|  | Satisfying | 30 (35) |
|  | Unknown | 2 (2) |
| Adjuvant therapy, n (%) | No therapy | 52 (60) |
|  | Vaginal brachytherapy | 10 (12) |
|  | External beam radiotherapy | 8 (9) |
|  | Chemotherapy | 11 (13) |
|  | External beam radiotherapy +<br>Chemotherapy | 4 (5) |
|  | Other | 1 (1) |
| Recurrence site, n (%) | No recurrence | 70 (81) |
|  | Vaginal | 0 (0) |
|  | Regional | 4 (5) |
|  | Distant | 12 (14) |

### Survival Outcomes and Recurrence Pattern

The 12-, 24-, and 36-month PFS rates were 91%, 78%, and 76%, respectively. Most recurrences occurred within 2 years after surgery, after which the survival curve showed relative stabilization (Fig. 1). Among the 16 patients who experienced recurrence, 12 (75%) developed distant metastases (Table 1).

**Fig. 1.**
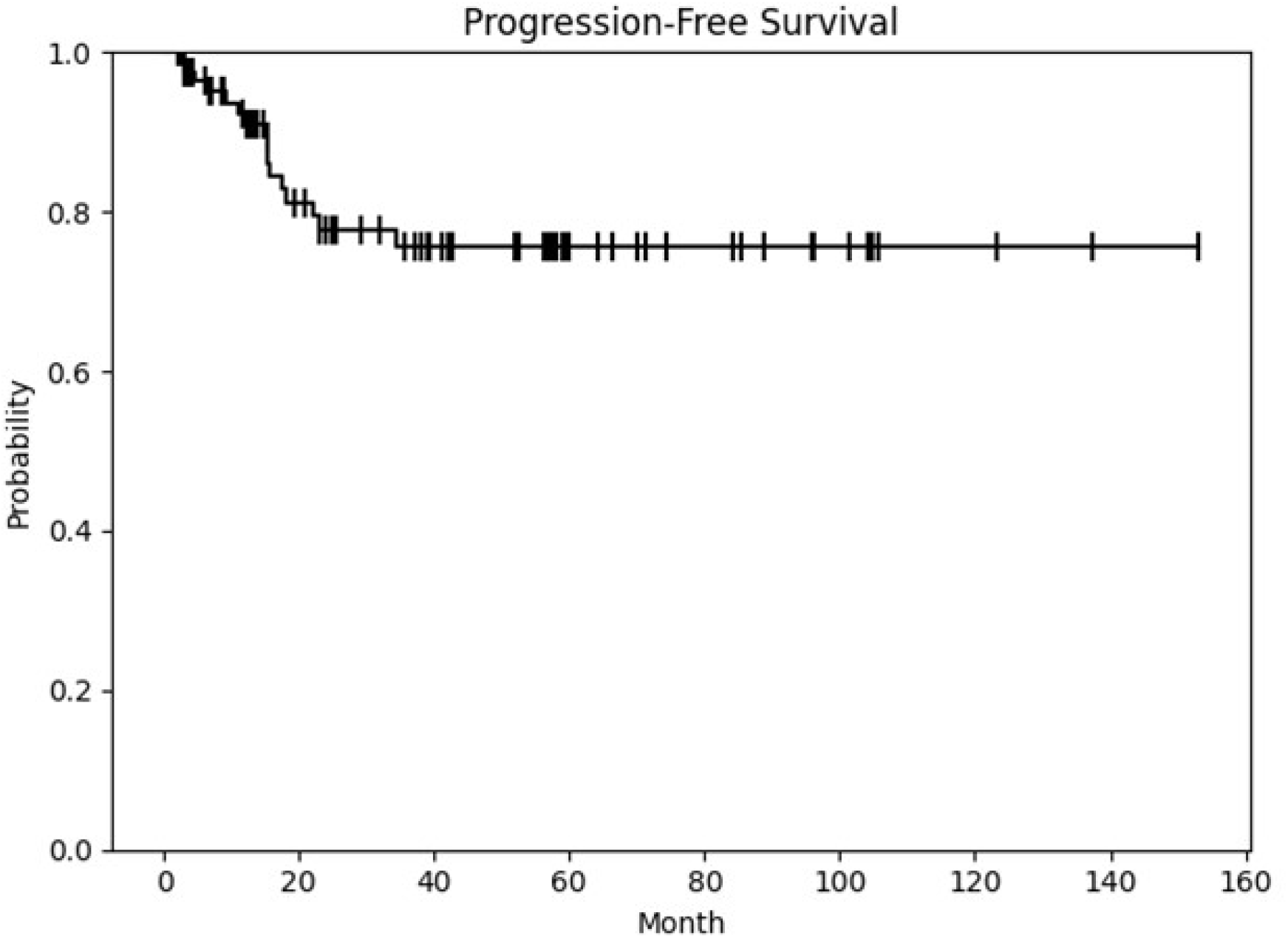
Kaplan–Meier curve of progression-free survival in the overall cohort.

### PFS According to Adjuvant Therapy

Patients who received chemotherapy alone demonstrated a numerically higher 2-year PFS compared with those who received external beam radiation therapy (EBRT) with or without chemotherapy (60% vs. 25%, log-rank p=0.10) (Fig. 2).

**Fig. 2.**
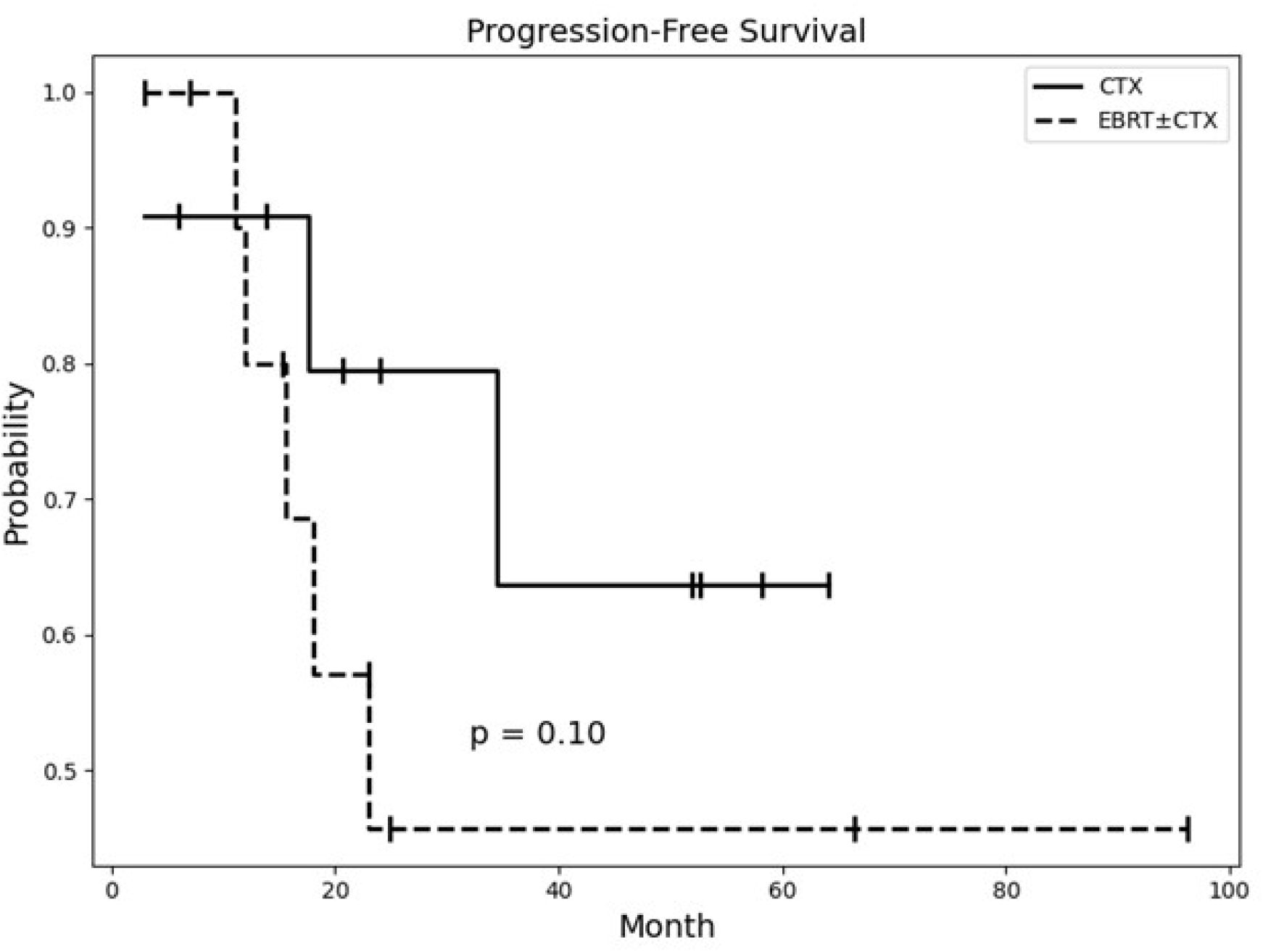
Kaplan–Meier curves of progression-free survival according to adjuvant therapy. CTX, chemotherapy; EBRT, external beam radiation therapy.

Baseline characteristics according to adjuvant therapy group are summarized in Table 2. Patients in the EBRT with or without chemotherapy group had significantly higher-risk features compared with those in the chemotherapy alone group, including older age (65 vs. 55 years, p<0.01), a higher proportion of the p53-overexpression subtype (75% vs. 9%, p<0.01), a higher rate of high-intermediate risk criteria (92% vs. 36%, p=0.01), and more advanced stage disease (Stage 2: 33% vs. 0%).

**Table 2.**
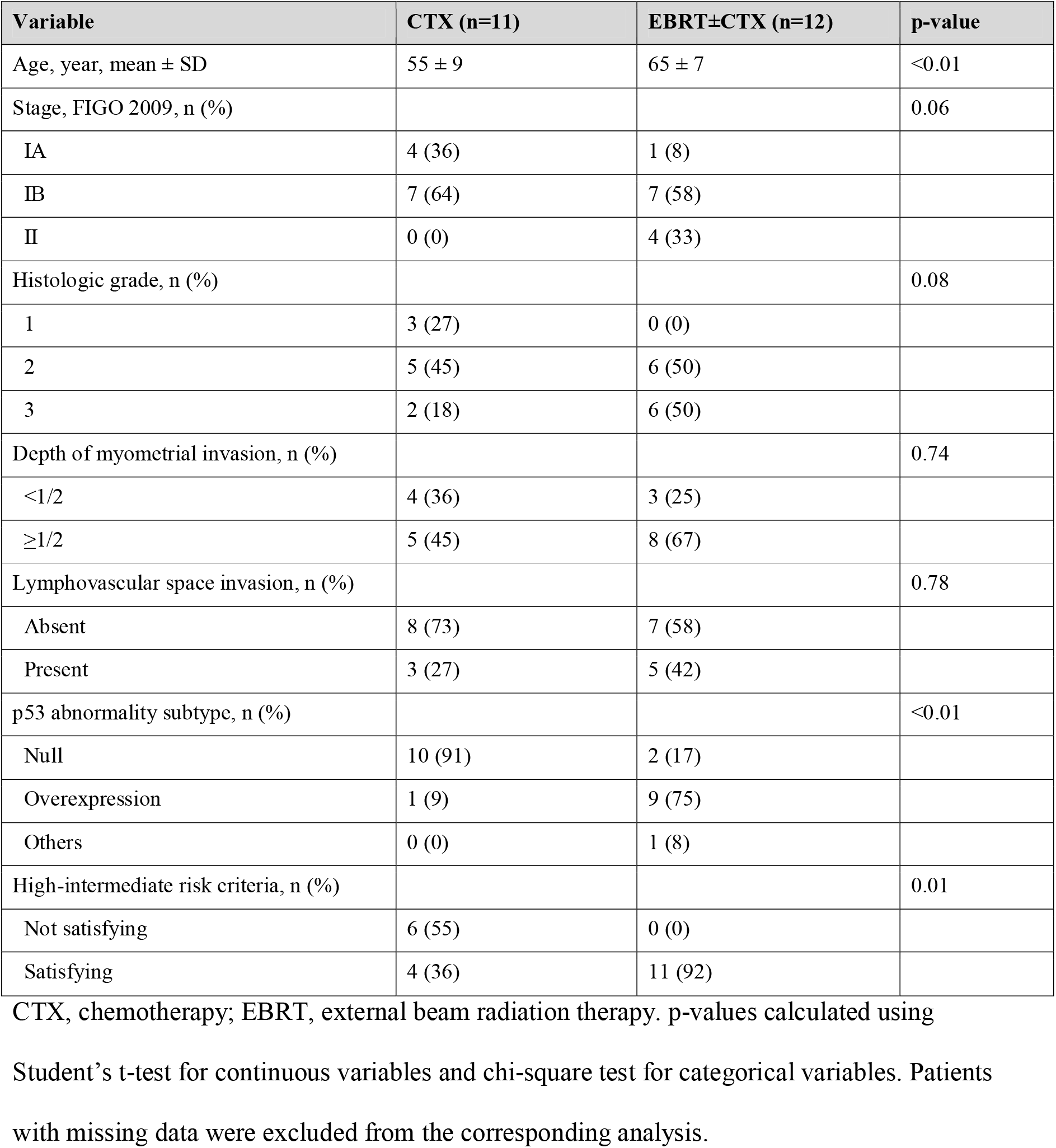
Baseline characteristics according to adjuvant therapy (chemotherapy alone vs. EBRT with or without chemotherapy)

| Variable | CTX (n=11) | EBRT±CTX (n=12) | p-value |
| --- | --- | --- | --- |
| Age, year, mean ± SD | 55 ± 9 | 65 ± 7 | <0.01 |
| Stage, FIGO 2009, n (%) |  |  | 0.06 |
| IA | 4 (36) | 1 (8) |  |
| IB | 7 (64) | 7 (58) |  |
| II | 0 (0) | 4 (33) |  |
| Histologic grade, n (%) |  |  | 0.08 |
| 1 | 3 (27) | 0 (0) |  |
| 2 | 5 (45) | 6 (50) |  |
| 3 | 2 (18) | 6 (50) |  |
| Depth of myometrial invasion, n (%) |  |  | 0.74 |
| <1/2 | 4 (36) | 3 (25) |  |
| ≥1/2 | 5 (45) | 8 (67) |  |
| Lymphovascular space invasion, n (%) |  |  | 0.78 |
| Absent | 8 (73) | 7 (58) |  |
| Present | 3 (27) | 5 (42) |  |
| p53 abnormality subtype, n (%) |  |  | <0.01 |
| Null | 10 (91) | 2 (17) |  |
| Overexpression | 1 (9) | 9 (75) |  |
| Others | 0 (0) | 1 (8) |  |
| High-intermediate risk criteria, n (%) |  |  | 0.01 |
| Not satisfying | 6 (55) | 0 (0) |  |
| Satisfying | 4 (36) | 11 (92) |  |

CTX, chemotherapy; EBRT, external beam radiation therapy. p-values calculated using
Student's t-test for continuous variables and chi-square test for categorical variables. Patients with missing data were excluded from the corresponding analysis.

### Prognostic Factors

In univariable analysis, the p53-overexpression pattern and negative ER expression were associated with worse PFS. Other clinicopathologic variables, including MMR status, were not significantly associated with PFS (Table 2). In multivariable analysis, the p53-overexpression pattern was independently associated with poorer PFS compared with the null pattern (HR 8.59, 95% CI 1.61–45.79, p=0.01) (Table 3). The Kaplan–Meier curves according to p53-abnormal subtype demonstrated clear separation between the overexpression and null patterns (Fig. 3).

**Table 3.** Association of variables with progression-free survival.

| Variable | HR (Univariable) | 95% CI | p-value | HR (Multivariable) | 95% CI | p-value |
| --- | --- | --- | --- | --- | --- | --- |
| Age |  |  |  |  |  |  |
| 35–55 | 1.00 | — | — | 1.00 | — | — |
| 56–61 | 0.50 | 0.16–1.53 | 0.22 | 0.54 | 0.07–4.14 | 0.55 |
| 62–84 | 0.42 | 0.11–1.59 | 0.20 | 0.11 | 0.00–2.96 | 0.19 |
| BMI |  |  |  |  |  |  |
| <25 | 1.00 | — | — | 1.00 | — | — |
| ≥25 | 0.89 | 0.33–2.37 | 0.82 | 0.90 | 0.21–3.88 | 0.89 |
| Menopause |  |  |  |  |  |  |
| Premenopause | 1.00 | — | — | 1.00 | — | — |
| Postmenopause | 0.44 | 0.16–1.17 | 0.10 | 0.14 | 0.02–1.08 | 0.06 |
| Stage, FIGO 2009 |  |  |  |  |  |  |
| IA | 1.00 | — | — | 1.00 | — | — |
| IB–II | 2.50 | 0.94–6.69 | 0.07 | NA | 0.00–inf | 1.00 |
| Histologic grade |  |  |  |  |  |  |
| 1 | 1.00 | — | — | 1.00 | — | — |
| 2 | 2.93 | 0.61–14.11 | 0.18 | 1.84 | 0.28–12.18 | 0.52 |
| 3 | 4.47 | 0.90–22.17 | 0.07 | 1.22 | 0.10–14.40 | 0.88 |
| Depth of myometrial invasion |  |  |  |  |  |  |
| <1/2 | 1.00 | — | — | 1.00 | — | — |
| ≥1/2 | 2.71 | 0.98–7.50 | 0.05 | 0.00 | 0.00–inf | 1.00 |
| Lymphovascular space invasion |  |  |  |  |  |  |
| Absent | 1.00 | — | — | 1.00 | — | — |
| Present | 2.18 | 0.76–6.26 | 0.15 | 0.90 | 0.20–4.16 | 0.90 |
| MMR status |  |  |  |  |  |  |
| Proficient | 1.00 | — | — | — | — | — |
| Deficient | 0.51 | 0.12–2.09 | 0.35 | — | — | — |
| p53 abnormality subtype |  |  |  |  |  |  |
| Null | 1.00 | — | — | 1.00 | — | — |
| Overexpression | 4.12 | 1.51–11.25 | <0.01 | 8.59 | 1.61–45.79 | 0.01 |
| ER expression |  |  |  |  |  |  |
| Positive | 1.00 | — | — | 1.00 | — | — |
| Negative | 3.06 | 1.02–9.14 | <0.05 | 2.35 | 0.51–10.96 | 0.28 |
| High-intermediate risk criteria |  |  |  |  |  |  |
| Not satisfying | 1.00 | — | — | — | — | — |
| Satisfying | 1.32 | 0.47–3.72 | 0.60 | — | — | — |

Patients with missing data for a given variable were excluded from the corresponding univariable analysis. Missing data included histologic grade (n=1), depth of myometrial invasion (n=3), MMR status (n=48), p53 staining subtype (n=4), ER expression (n=11), and high-intermediate risk status (n=2). Multivariable analysis was performed using cases with complete data for all included variables (n=67).

**Fig. 3.**
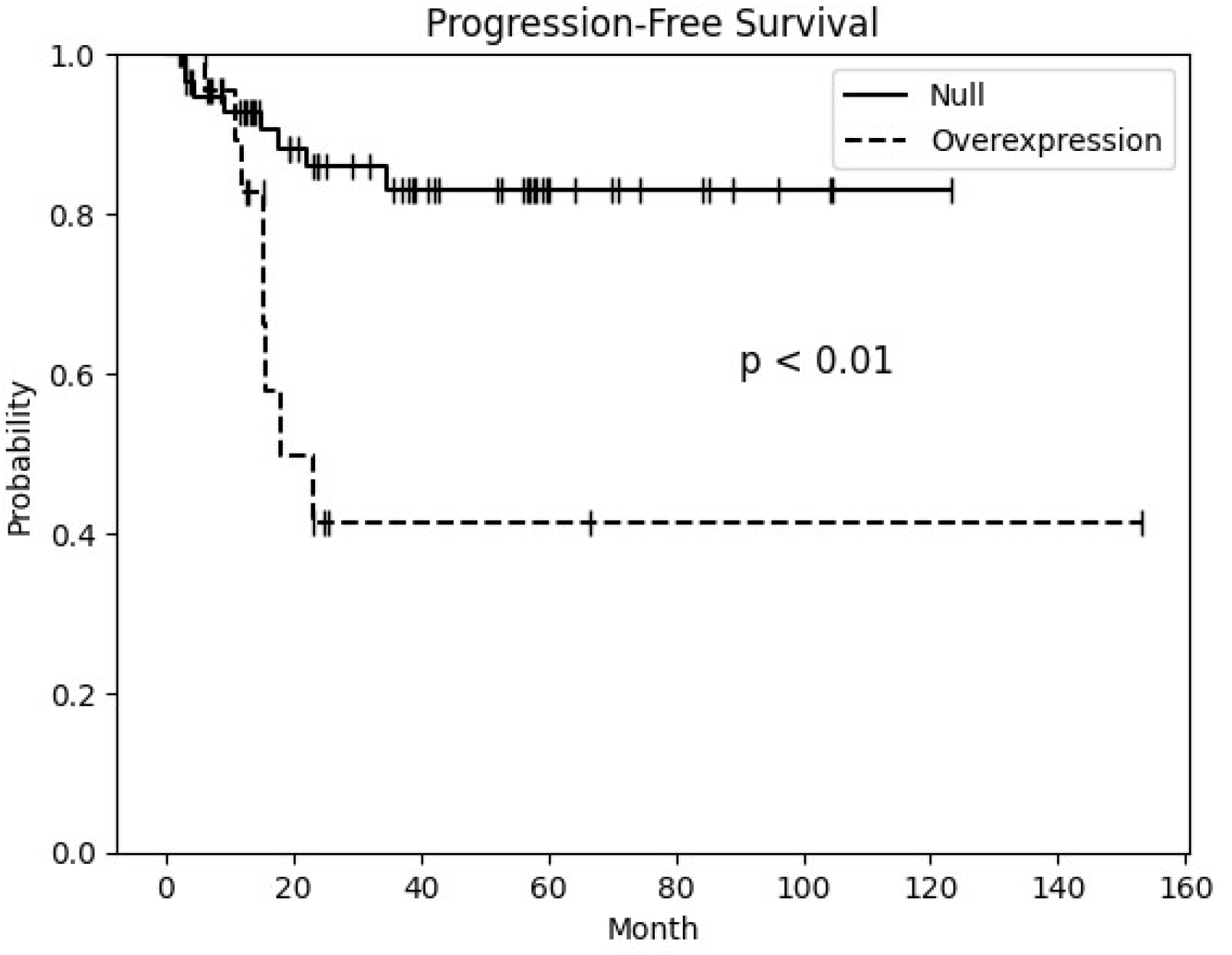
Unadjusted Kaplan–Meier curves of progression-free survival according to p53 abnormality subtype.

Notable differences in clinicopathologic characteristics and recurrence patterns were observed between p53 abnormality subtypes (Table 4). Patients with the p53-overexpression pattern were significantly older (62 vs. 57 years, p=0.03) and more frequently had higher histologic grade (grade 3: 57% vs. 16%, p<0.01) and high-intermediate risk criteria (59% vs. 28%, p=0.02) compared with those with the null pattern. Distant metastases were more frequent in the p53-overexpression group, whereas regional recurrence occurred only in the null-type group.

**Table 4.** Clinicopathologic characteristics, adjuvant therapy, and recurrence pattern according to p53 abnormality subtype.

| Variable | Null (n=59) | Overexpression (n=23) | p-value |
| --- | --- | --- | --- |
| Age, year, mean ± SD | 57 ± 9 | 62 ± 12 | 0.03 |
| BMI, mean ± SD | 26 ± 5 | 25 ± 5 | 0.29 |
| Menopause, n (%) |  |  | 0.18 |
| Premenopause | 18 (31) | 3 (13) |  |
| Postmenopause | 41 (69) | 20 (87) |  |
| Stage, FIGO 2009, n (%) |  |  | 0.39 |
| IA | 43 (73) | 14 (61) |  |
| IB | 13 (22) | 6 (26) |  |
| II | 3 (5) | 3 (13) |  |
| Histologic grade, n (%) |  |  | <0.01 |
| 1 | 27 (47) | 1 (4) |  |
| 2 | 22 (38) | 9 (39) |  |
| 3 | 9 (16) | 13 (57) |  |
| Depth of myometrial invasion, n (%) |  |  | 0.27 |
| <1/2 | 45 (79) | 14 (64) |  |
| ≥1/2 | 12 (21) | 8 (36) | |
| Lymphovascular space invasion, n (%) |  |  | 0.09 |
| Absent | 52 (88) | 16 (70) |  |
| Present | 7 (12) | 7 (30) |  |
| High-intermediate risk criteria, n (%) |  |  | 0.02 |
| Not satisfying | 42 (72) | 9 (41) |  |
| Satisfying | 16 (28) | 13 (59) |  |
| Adjuvant therapy, n (%) |  |  |  |
| No therapy | 40 (68) | 9 (39) | NA |
| Vaginal brachytherapy | 7 (12) | 3 (13) |  |
| External beam radiotherapy | 1 (2) | 6 (26) |  |
| Chemotherapy | 10 (17) | 1 (4) |  |
| External beam radiotherapy + Chemotherapy | 1 (2) | 3 (13) |  |
| Other | 0 (0) | 1 (4) |  |
| Type of recurrence, n (%) |  |  |  |
| No recurrence | 51 (86) | 15 (65) | NA |
| Vaginal | 0 (0) | 0 (0) |  |
| Regional | 4 (7) | 0 (0) |  |
| Distant | 4 (7) | 8 (35) |  |

p-values calculated using Student's t-test for continuous variables and chi-square test for categorical variables. Patients with missing data were excluded from the corresponding analysis.

## Discussion

Although p53 abnormalities are more commonly associated with non-endometrioid histologic subtypes, they can also occur in endometrioid endometrial cancers and have been shown to confer a distinctly aggressive clinical behavior even within this histologic subtype. For example, Jamieson et al. examined a large international cohort of stage I low-grade p53-abnormal endometrioid endometrial carcinomas and reported significantly worse disease-specific survival compared with p53 wild-type counterparts, with a recurrence rate substantially higher than expected for conventional low-grade endometrioid tumors [10]. Similarly, Chang et al. reported that among patients with stage I, grade 1 or 2 endometrioid carcinoma, those with abnormal p53 expression had a recurrence rate of 19.2%, whereas no recurrences were observed in the p53-normal group [11]. Collectively, these findings support the notion that p53 abnormality increases the biological risk of endometrioid early-stage endometrial cancer beyond what is predicted by conventional clinicopathologic factors alone. Our findings are consistent with this body of evidence, demonstrating a 3-year PFS of 76% and a high rate of distant recurrence in this population.

Although only patients with early-stage disease were included, the 3-year PFS rate in our cohort was 76%, which is lower than expected for conventional endometrioid early-stage endometrial cancer. These findings suggest that patients with a p53-abnormal pattern may represent a biologically aggressive subgroup despite early-stage presentation. Notably, more than half of the patients in this cohort did not receive adjuvant therapy. This may reflect treatment decisions based primarily on traditional clinicopathologic risk factors rather than molecular classification. Our findings support the consideration of molecular risk stratification in guiding adjuvant treatment decisions.

Among the 16 recurrences observed, 12 were distant metastases, indicating that systemic failure was more frequent than locoregional relapse. Patients who received chemotherapy alone demonstrated a numerically higher 2-year PFS compared with those who received EBRT with or without chemotherapy (60% vs. 25%, log-rank p=0.10); however, this difference did not reach statistical significance. As shown in Table 2, patients in the EBRT with or without chemotherapy group had significantly higher baseline risk profiles, suggesting that the numerically lower PFS in this group likely reflects selection bias rather than a true inferiority of radiation-based treatment. Therefore, the results should not be construed as evidence that chemotherapy is superior to EBRT-based regimens. Nevertheless, the predominance of distant recurrence in this cohort suggests a potential role for systemic therapy in this population, and prospective studies with balanced risk profiles are needed to draw definitive conclusions.

We also observed that the p53-overexpression pattern was independently associated with poorer PFS compared with the null pattern. Although the number of events was limited and the confidence interval was wide, this finding raises the possibility that subtypes of p53 abnormality may have distinct prognostic implications. Further validation in independent cohorts is warranted.

As shown in Table 4, patients with the p53-overexpression pattern had significantly higher-risk clinicopathologic features compared with those with the null pattern, including older age, higher histologic grade, and more frequent high-intermediate risk criteria. These differences suggest that the higher rate of distant recurrence observed in the overexpression group may not be attributable solely to p53 abnormality, but may also reflect the co-occurrence of other established risk factors. Given the limited sample size and the small number of events, a fully adjusted multivariable analysis of distant recurrence was not feasible in this study. These findings underscore the importance of integrating both molecular and clinicopathologic risk factors when assessing recurrence risk in this population, and highlight the need for prospective studies with comprehensive molecular and clinicopathologic characterization.

Previous studies have reported favorable outcomes in patients with dMMR tumors, suggesting that dMMR may represent a less aggressive molecular subgroup [12]. In contrast, MMR status was not associated with PFS in our study. This discrepancy may be explained by the limited sample size, incomplete MMR data, and the absence of POLE mutation testing, which may have led to misclassification within molecular subgroups.

We excluded tumors without myometrial invasion because the benefit of adjuvant therapy in this setting remains controversial. In this context, parallels may be drawn with uterine serous carcinoma, which is generally classified as high risk; however, the optimal adjuvant strategy for stage IA tumors confined to the endometrium remains debated. Some retrospective studies suggest favorable outcomes with observation after comprehensive staging, whereas others report a clinically meaningful risk of distant recurrence, supporting consideration of systemic chemotherapy [13].

This study has several limitations, including its retrospective design, limited number of events, and potential selection bias. Molecular classification was incomplete in a substantial proportion of patients: MMR status was unavailable in 56% of patients, and POLE mutational status was not assessed in any patient. According to the hierarchical molecular classification algorithm — in which POLE-ultramutated tumors are prioritized first, followed by dMMR, then p53-abnormal, and finally NSMP — the absence of POLE and complete MMR data means that some patients classified as p53-abnormal in this study may harbor concurrent POLE mutations or dMMR status, which carry distinct and generally more favorable prognostic implications. This incomplete molecular characterization may have introduced misclassification bias, potentially affecting the observed survival outcomes. Furthermore, complete chemotherapy data, including regimen details, number of cycles, and treatment intensity, were not systematically collected for all patients. Although paclitaxel plus carboplatin was used in patients receiving chemotherapy alone and cisplatin-based regimens in those receiving EBRT combined with chemotherapy, the absence of complete treatment data limits the interpretation of chemotherapy-related outcomes. Therefore, the results should be interpreted cautiously.

In conclusion, patients with endometrioid early-stage endometrial cancer exhibiting a p53-abnormal pattern showed relatively unfavorable PFS and a high rate of distant recurrence. Although the survival difference did not reach statistical significance, adjuvant chemotherapy may be considered in selected patients. Prospective studies are needed to clarify the optimal adjuvant strategy and the prognostic significance of p53 abnormality subtypes.

## Data Availability

All data produced in the present study are available upon reasonable request to the authors

## Acknowledgments

None.

